# Rule of thumb in human intelligence for assessing the COVID-19 outbreak in Japan

**DOI:** 10.1101/2021.01.20.21250204

**Authors:** Junko Kurita, Tamie Sugawara, Yasushi Ohkusa

## Abstract

**Background:** The COVID-19 outbreak in Japan exhibited its third peak at the end of 2020. Mathematical modelling and developed AI cannot explain several peaks in a single year.

**Object:** This study was conducted to evaluate a rule of thumb for prediction from past wave experiences.

**Method:** We rescaled the number of newly infected patients as 100% at the peak and checked similarities among waves. Then we extrapolated the courses of the third and later waves.

**Results:** Results show some similarity around the second and the third waves. Based on this similarity, we expected the bottom of the third wave will show 2131 newly positive patients including asymptomatic patients at around the end of February, 2021.

**Discussion and Conclusion:** We can infer the course of the third wave from similarity with the second wave. Mathematical modelling has been unable to do it, even when AI was used for prediction. Performance of the rule of thumb used with human intelligence might be superior to that of AI under these circumstances.

## Introduction

Three waves of the COVID-19 outbreak have affected Japan since the emergence of COVID-19 in Wuhan, China in December, 2019. The third wave peaked at the end of 2020, then declined as of January 17, 2021 (Figure 1).

**Figure 1:**
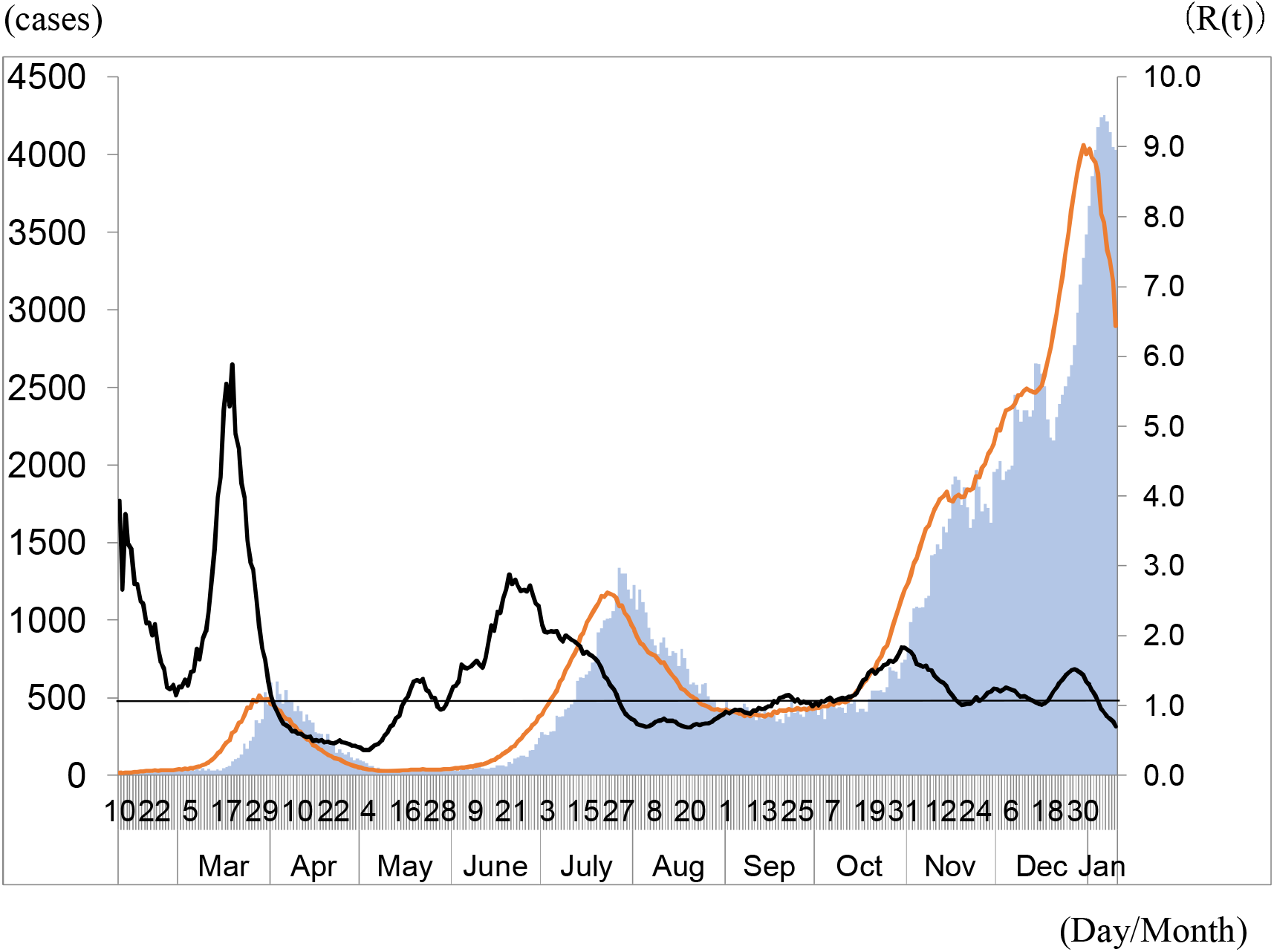
Epidemic curve, with newly infected symptomatic patients and R(t) in Japan. Note: Bars represent the epidemic curve, the number of patients showing new onset each day. The red line represents the number of newly infected symptomatic patients. The black line shows R(t).

Prediction of the course of the outbreak is expected to be most important for countermeasures and preparations for it. Some mathematical modelling [1–3] including developed AI [4] has been proposed. Nevertheless, it principally cannot explain several waves in a single year.

By contrast, a “rule of thumb” used with human intelligence reflects information, knowledge and experience obtained in the past. Such a rule can predict outcomes of the near future with no rational process necessary. In this sense, a rule of thumb used with human intelligence has some probability of surpassing the performance of an AI. For the present study, we apply a rule of thumb with human intelligence to prediction of the COVID-19 outbreak in Japan.

## Method

The numbers of symptomatic patients reported by the Ministry of Health, Labour and Welfare (MHLW) for January 14 – November 30, published [5] as of December 25 were used. Some patients were excluded from data for Japan: those presumed to be persons infected abroad or infected as Diamond Princess passengers. Those patients were presumed not to represent community-acquired infection in Japan. For onset dates of some symptomatic patients that were unknown, we estimated their onset date from an empirical distribution with duration extending from onset to the report date among patients for whom the onset date had been reported.

The following procedure is similar to that used for our earlier research [6,7]. First, we estimated the onset date of patients for whom onset dates were not reported. Letting *f*(*k*) represent this empirical distribution and letting *N*_*t*_ denote the number of patients for whom onset dates were not available published at date *t*, then the number of patients for whom the onset date was known is *t*-1. The number of patients for whom onset dates were unavailable was estimated as *f*(1)*N*_*t*_. Similarly, the number of patients with onset date *t*-2 and for whom onset dates were not available was estimated as *f*(2)*N*_*t*_. Therefore, the total number of patients for whom the onset date was not available, given an onset date of *s*, was estimated as Σ_*k*=1_ *f*(*k*)*N*_*s*_+*k* for the long duration extending from *s*.

Moreover, the reporting delay for published data from MHLW might be considerable. In other words, if *s*+*k* is larger than that in the current period *t*, then *s*+*k* represents the future for period *t*. For that reason, *Ns*+*k* is not observable. Such a reporting delay leads to underestimation of the number of patients. For that reason, it must be adjusted as 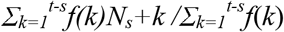. Similarly, patients for whom the onset dates were available are expected to be affected by the reporting delay. Therefore, we have 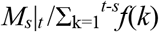 where *M*_*s*_|_*t*_ represents the reported number of patients for whom onset dates were within period *s*, extending until the current period *t*.

We defined R(t) as the number of infected patients on day *t* divided by the number of patients who were presumed to be infectious. The number of infected patients was calculated from the epidemic curve by the onset date using a distribution of the incubation period. The distribution of infectiousness in symptomatic and asymptomatic cases was assumed to be 30% on the onset day, 20% on the following day, and 10% for the subsequent five days [8].

We rescaled the newly infected symptomatic patients as percentage of the peak in each wave. The time horizon was also adjusted as peak date was 0. Then we checked some similarity among waves. If we found some similarity, then we extrapolated it to the third and later waves. The study period extended from February 10, 2020 to January 10 as of January 17, 2021.

## Ethical considerations

All information used for this study has been published elsewhere [5]. There is therefore no ethical issue related to its use for this study.

## Results

From Figure 1, we found the first peak as March 28. The second was July 28. The third one was December 28. We defined the first wave as lasting until May 15. The second wave extended until September 15, 2020. When we rescaled the newly infected symptomatic cases and adjusted the date, the three lines were aligned as portrayed in Figure 2. Results showed that the second and third wave were almost identical around the peak. Therefore, we expected that the line in the third wave declined following the line in the second wave. Then it can be expected to reach the bottom about 50 days after the peak. It was expected to be 35% of the peak at the bottom.

**Figure 2:**
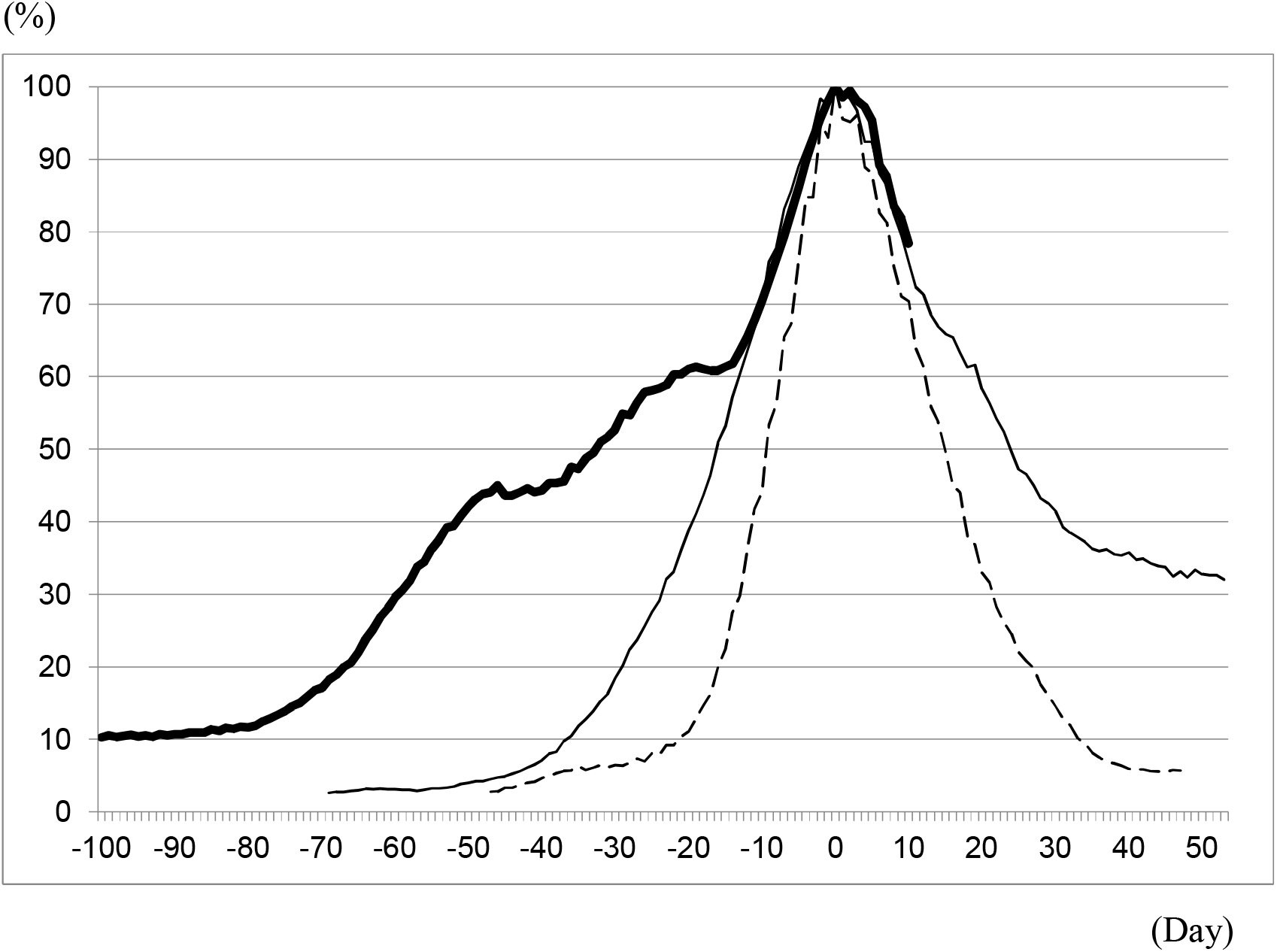
Percentage of newly infected symptomatic patients to the respective peaks of three waves. Note: The broken line represents the percentage of the newly infected symptomatic patients to its peak in the first wave. The thin line represents those in the second wave. The bold line represents those in the third wave. Day 0 was adjusted as the peak date in each wave.

The lead time from the bottom to the peak in the following wave was 70 days in the second wave and 106 days in the third wave. That period corresponds to the end of April extending to the beginning of June 2021.

## Discussion

We expected the bottom of the third wave as around the end of February and starting the fourth wave at that time. At that time, the newly infected symptomatic patients are expected to be about 35% of the peak, which was approximately 1,400 symptomatic patients. Because asymptomatic patients were one third of all tested-positive patients, one can infer 2,100 positive patients.

Some researchers provided their predictions and policy evaluations [9]. However, the baseline scenario with no aggressive policy such as requirement of voluntary restriction for going out or shorter business hours in restaurants was increasing monotonically for more than three months. Therefore, they certainly failed to incorporate the fact that the peak of the third wave had already been passed at the end of last year, 2020.

Google provided AI prediction in Japan [10], as shown in Figure 3. Their prediction was for the number of newly tested-positive patients each day, which is not the same as newly infected symptomatic patients inferred in the present study. The Google AI clearly predicted a monotonic increase until mid-February. Therefore, it is clearly inconsistent with our prediction based on our rule of thumb. It remains unclear which is the better prediction at this time. However, as shown in Figure 3, the number of newly tested-positive patients also appears to be declining in January, although the day of the week affects the result heavily. Tentatively, as of January 17, 2021, the rule of thumb in human intelligence appears to be better than AI.

**Figure 3:**
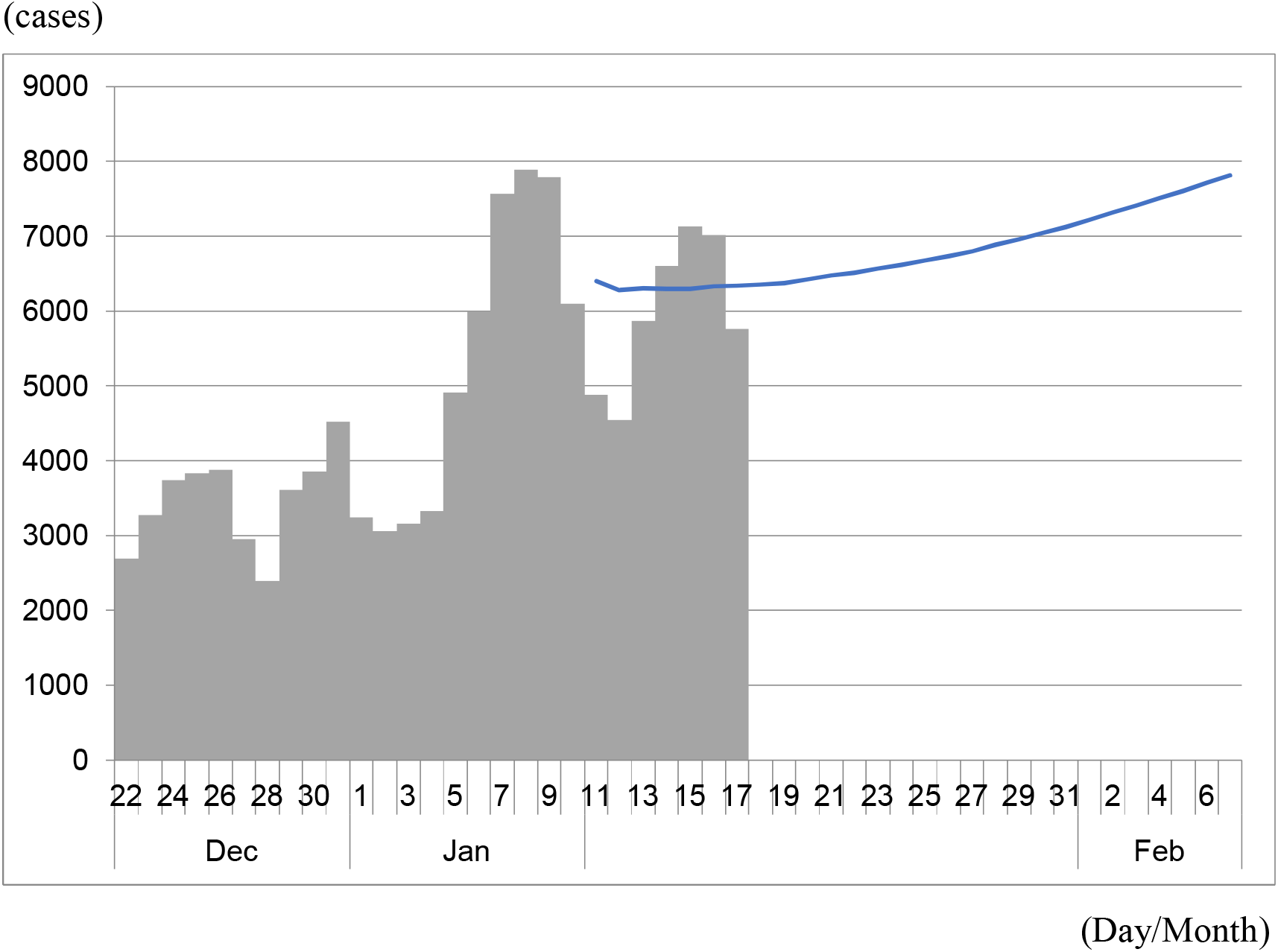
Reported newly tested-positive patients and its value predicted by Google AI from January 11 through February 7, 2021 as of January 10, 2021. Note: Bars represent the reported newly test positive patients including asymptomatic patients in each day. The line indicates its prediction by Google AI as of January 10, 2021.

In principle, AI presents greater advantages at finding a rule of thumb than human intelligence because it can use greater amounts of information and knowledge on many dimensions. It can evaluate any association at lower cost than human intelligence can. An AI that is able to use a rule of thumb can be expected to be more powerful than a rule of thumb with human intelligence. However, at least to date, AIs have been restricted to using mathematical modelling. They have been abandoned for use in ascertaining some rule of thumb. Therefore, a rule of thumb used with human intelligence might be more powerful than an AI restricted to mathematical modelling. In other words, mathematical modelling appears to be insufficient for predicting the course of a COVID-19 outbreak.

For the present study, the authors inferred rule of thumb from the prior two waves. However, only two waves can be expected to provide insufficient information for precise prediction. Improving and accumulating experience of waves of infection is expected to improve the precision of the rules of thumb.

Especially, by definition, a rule of thumb can be expected to be very weak for new situations that one has not experienced, such as vaccine initiation or virus mutation. Mathematical modelling might resurge in importance for assessing such situations.

## Conclusion

We predicted the bottom of the third wave to occur around the end of February, with approximately 1,400 symptomatic patients at that time. A rule of thumb used with human intelligence might be better than AI using the mathematical model.

The present study is based on the authors’ opinions: it does not reflect any stance or policy of their professionally affiliated bodies.

## Data Availability

Japan: COVID-19 Public Forecasts

https://datastudio.google.com/u/0/reporting/8224d512-a76e-4d38-91c1-935ba119eb8f/page/ncZpB

## Acknowledgments

We acknowledge the great efforts of all staff at public health centers, medical institutions, and other facilities who are fighting the spread and destruction associated with COVID-19.

## Ethical considerations

All information used for this study was collected under the Law of Infection Control, Japan and published data was used. There is therefore no ethical issue related to this study.

## Reference

1. Kissler SM, Tedijanto C, Goldstein E, Grad YH, Lipsitch M. Projecting the Transmission Dynamics of SARS-CoV-2 Through the Postpandemic Period. Science 2020; 368:860–8.

2. Sugishita Y, Kurita J, Sugawara T, Ohkusa Y. Effects of voluntary event cancellation and school closure as countermeasures against COVID-19 outbreak in Japan. Plos one 2020. https://doi.org/10.1371/journal.pone.0239455

3. Kurita J, Sugawara T, Ohkusa Y. Estimated effectiveness of school closure and voluntary event cancellation as COVID-19 countermeasures in Japan. J Infect Chemother 2021, 27:62–4. DOI: 10.1016/j.jiac.2020.08.012. Epub 2020 Aug 19.

4. Arik S, Li C, Yoon J, Sinha R, Epshteyn A, Le LT, Menon V, Singh S, Zhang L, Nikoltchev M, Sonthalia YK, Nakhost H, Kanal E, Pfister T. Interpretable Sequence Learning for COVID-19 Forecasting. NeurIPS 2020.

5. https://research.google/pubs/pub49500/ Japan Times. Tokyo governor urges people to stay indoors over weekend as virus cases spike https://www.japantimes.co.jp/news/2020/03/25/national/science-health/tokyo-logs-40-coronavirus-cases/#.Xr4TV2eP604 [accessed on May 14, 2020

6. Kurita J, Sugawara T, Ohkusa Y. Estimated effectiveness of school closure and voluntary event cancellation as COVID-19 countermeasures in Japan. J Infect Chemother 2021, 27:62–4. DOI: 10.1016/j.jiac.2020.08.012. Epub 2020 Aug 19.

7. Sugishita Y, Kurita J, Sugawara T, Ohkusa Y. Effects of voluntary event cancellation and school closure as countermeasures against COVID-19 outbreak in Japan. Plos one 2020. https://doi.org/10.1371/journal.pone.0239455

8. Kimball A, Hatfield KM, Arons M, James A, Taylor J, Spicer K, Bardossy AC, Oakley LP, Tanwar S, Chisty Z, Bell JM, Methner M, Harney J, Jacobs JR, Carlson CM, McLaughlin HP, Stone N, Clark S, Brostrom-Smith C, Page LC, Kay M, Lewis J, Russell D, Hiatt B, Gant J, Duchin JS, Clark TA, Honein MA, Reddy SC, Jernigan JA; Public Health Seattle & King County; CDC COVID-19 Investigation Team. Asymptomatic and Presymptomatic SARS-CoV-2 Infections in Residents of a Long-Term Care Skilled Nursing Facility - King County, Washington, March 2020. Morb Mortal Wkly Rep. 2020 Apr 3;69(13):377–381. DOI: 10.15585/mmwr.mm6913e1.

9. Nippon Housou Kyoukai. Professor simulates virus spread in Tokyo. https://www3.nhk.or.jp/nhkworld/en/news/20210105_39/ [accessed on 21 January, 2021]

10. https://datastudio.google.com/reporting/8224d512-a76e-4d38-91c1-935ba119eb8f (in Japanese) [accessed on 21 January, 2021]

